# A multivariate genetic analysis of environmental sensitivity, anxiety sensitivity and reported life events in adolescents

**DOI:** 10.1101/2022.05.24.22275523

**Authors:** Alicia J. Peel, Olakunle Oginni, Elham Assary, Georgina Krebs, Celestine Lockhart, Thomas McGregor, Elisavet Palaiologou, Angelia Ronald, Andrea Danese, Thalia C. Eley

## Abstract

**Background:** Despite being considered a measure of environmental risk, reported life events are partly heritable. One mechanism that may contribute to their heritability is genetic influences on sensitivity. These sensitivity biases can relate to how individuals process the contextual aspects of their environment (environmental sensitivity) or how they interpret their own physical and emotional responses (anxiety sensitivity). The aim of this study was to explore the genetic and environmental overlap between self-reported life events and measures of sensitivity.

**Methods:** At age 17, individuals (N = 2,939) from the Twins Early Development Study (TEDS) completed measures of environmental sensitivity (Highly Sensitive Child Scale) and anxiety sensitivity (Children’s Anxiety Sensitivity Index), as well as reporting on their experience of 20 recent life events. Using multivariate Cholesky decomposition models, we investigated the shared genetic and environmental influences on the associations between these measures of sensitivity and the number of reported life events, as well as both negative and positive ratings of life events.

**Results:** The majority of the associations between anxiety sensitivity, environmental sensitivity and reported life events were explained by shared genetic influences (59%-75%), with the remainder explained by non-shared environmental influences (25%-41%). Environmental sensitivity showed comparable genetic correlations with both negative and positive ratings of life events (r_A_ = .21 and .15), anxiety sensitivity only showed a significant genetic correlation with negative ratings of life events (r_A_ = .33). Approximately 10% of the genetic influences on reported life events were accounted for by genetic influences shared with anxiety sensitivity and environmental sensitivity.

**Conclusions:** A proportion of the heritable component of reported life events is captured by measures of sensitivity. Differences in how individuals process the contextual aspects of the environment or interpret their own physical and emotional response to environmental stimuli may be one mechanism through which genetic liability influences the subjective experience of life events.

## Introduction

Adverse life events are associated with numerous poor outcomes, including emotions (Eley & Stevenson, 2000), behavioural (Flouri & Mavroveli, 2013) and thought problems (Shakoor et al., 2016). The period of adolescence is central to understanding the relationships between these events and outcomes. During this time, exposure to stressors such as puberty, social pressures and academic milestones significantly increases (Grant et al., 2004) and the brain is differentially susceptible to the adverse effects of these stressors (Lupien et al., 2009). Furthermore, the majority of mental illnesses emerge shortly after this period, in early adulthood (Solmi et al., 2021).

Despite being viewed as a measure of environmental risk, major life events have been found to have a significant genetic component. In a meta-analysis of 55 twin studies, self-reported stressful life events had an average heritability estimate of 29% (Kendler & Baker, 2007). Similarly, results from genome-wide association studies (GWAS) indicate that ∼30% of variance in self-reported stressful life events is explained by common genetic variants (Power et al., 2013). The identification of a heritable component for these environmental measures suggests that aspects of the experience of life events are not independent from genetic influences. This points to the possibility of two potential mechanisms of gene-environment interplay that may underlie the pathway from genes to experience.

The first of these potential pathways is gene-environment correlation (rGE), whereby a person’s genotype influences the types of environments that they are exposed to (Jaffee & Price, 2007). This can occur passively through effects on upbringing (Dale et al., 2015), actively or evocatively, for example, through behaviour (McAdams et al., 2013; Uliaszek et al., 2010) or personality traits (Hicks et al., 2013). In the case of life events, genetic predisposition for certain traits may make individuals more likely to encounter either adverse or positive environments. For example, negative life events in adolescence show genetic overlap with traits such as oppositionality, delinquency, physical aggression, depression, anxiety and neuroticism (McAdams et al., 2013). Positive events share genetic influences with wellbeing traits, such as ambition and hopefulness (Wootton et al., 2017). This suggests that part of the association between negative life events and poor outcomes could arise due to genetic confounding, whereby the same genes influence the likelihood of both experiencing an event and of developing a mental health disorder. In this framework, genetic predisposition to certain traits influences the *occurrence* of certain environments.

A second possibility is gene-environment interaction (GxE), which describes how the effects of genetics can vary depending on environmental exposures (Hunter, 2005). One example of how GxE may operate is through genetic influences on individual sensitivity to environmental experiences (Thapar et al., 2007). That is, some individuals are more susceptible to certain environments, and may be predisposed to experiencing adverse outcomes following exposure. In this framework, genes do not influence the *occurrence* of an event, but rather the *effect* of that event on the individual. This may explain why there is considerable variation in individuals’ responses to the same environments, with many who report negative life events displaying resilience and no adverse outcomes (Kim-Cohen et al., 2004). Gene-environment correlation and interaction are not mutually exclusive, and it is likely that both influence the experience of life events. However, significant gaps remain in our understanding of GxE, as the majority of past research has focussed on specific disorders using candidate genes, a methodology now superseded by genome-wide methods (Leighton et al., 2017). For example, the interaction between the serotonin transporter (5HTTLPR) short allele and life events has been widely researched in relation to the aetiology of depression (Caspi et al., 2003; Culverhouse et al., 2018; Munafò et al., 2009; Uher & McGuffin, 2010). Taking a broader genetic and transdiagnostic approach to exploring GxE may have greater potential to elucidate the genetic basis of general environmental sensitivity, rather than just the components that are shared with certain disorders.

In recent years, the understanding of GxE has shifted from focusing only on “vulnerability” to negative experiences towards broader “sensitivity” to both negative and positive contexts (Leighton et al., 2017). Sensitivity to contextual aspects of the environment, termed differential susceptibility, is proposed to moderate both the adverse effects of negative experiences as well as the tendency to benefit from positive and supportive environments (Belsky & Pluess, 2009). Measures that capture the thoughts and behaviours of sensitive individuals, such as the Highly Sensitive Child Scale, can be used to directly assess individual differences in broad environmental sensitivity (Pluess et al., 2018). In adolescent twins, genetic influences accounted for 47% of the variance in environmental sensitivity as measured through the Highly Sensitive Child Scale, and were found to overlap with influences on the personality traits of neuroticism and extraversion (Assary et al., 2021). In support of the differential susceptibility model, the genetic basis of this measure was found to consist of three heritable components: one reflecting general sensitivity, a second capturing the specific genetic influences that underlie reactivity to adversity, and a third that is relevant to responses to positive aspects of the environment (Assary et al., 2021).

Another form of differential sensitivity to experiences is the extent to which individuals negatively interpret their own physical and emotional responses to the environment. One such bias is anxiety sensitivity, the enhanced awareness of the symptoms of anxiety, such as heart palpitations or worry, and tendency to perceive these as being harmful (Taylor, 2014). Heritability estimates for anxiety sensitivity range from 37% in children (Eley et al., 2007) to 45-50% in adolescents and adults (Stein et al., 1999; Zavos et al., 2010). Anxiety sensitivity is also found to share genetic influences with several psychological traits, such as depression, anxiety and panic (Eley et al., 2007; Waszczuk et al., 2015; Zavos et al., 2010; Zavos, Rijsdijk, et al., 2012).

Phenotypically, both environmental sensitivity and anxiety sensitivity have been associated with stronger responses to major life events (Iimura, 2021; McLaughlin & Hatzenbuehler, 2009; Zavos, Wong, et al., 2012). High environmental sensitivity has been associated with both increases in mood and wellbeing following positive life events (Iimura, 2021) and decreases in wellbeing following exposure to stressful environments (Pluess et al., 2020). Notably, high environmental sensitivity was not associated with wellbeing at baseline, indicating that it does not reflect a generally lower emotional state, but rather increased sensitivity to the context of the environment (Pluess et al., 2020). Anxiety sensitivity has been shown to mediate the association between stressful life events and later anxiety symptoms in adolescence (McLaughlin & Hatzenbuehler, 2009), and between parent psychopathology and later anxiety in children (Drake & Kearney, 2008). These findings suggest that environmental sensitivity may be related to amplification of the impact of both positive and negative life events, while anxiety sensitivity primarily relates to negative interpretation and may exacerbate the impact of negative life events specifically.

However, it remains unclear how sensitivity to the context of the environment (environmental sensitivity) and sensitivity to emotional and physical reactions to the environment (anxiety sensitivity) are related, and whether they share a genetic basis. Additionally, there is limited understanding of the extent to which these traits explain the genetic underpinnings of reported life events. This knowledge would contribute to our understanding of what is captured by the heritable component of measures of environmental risk and the role of differential sensitivity in the self-reporting of environmental experiences.

### Aims

This study aimed to:

1. Investigate the shared genetic and environmental influences on the associations between environmental sensitivity, anxiety sensitivity and reported life events.
2. Explore whether these associations differ for life events rated as having negative valence as compared to those rated as having positive valence.
3. Quantify the proportion of genetic and environmental influences on reported life events that is shared with environmental sensitivity and anxiety sensitivity.

## Methods

All analyses were pre-registered on the Open Science Framework (OSF) prior to accessing the data (https://osf.io/haud4/). Custom code for these analyses is available on the OSF website.

### Sample

Data for this study were drawn from the Twins Early Development Study (TEDS), a study of over 15,000 twin pairs born in England and Wales between 1994-1996, identified through birth records (Rimfeld et al., 2019). The twins have been followed longitudinally throughout childhood into adulthood. At several time points, cognitive, emotional and behavioural data have been collected from the twins, their parents and teachers.

Between ages 15-17 years, 5,163 families returned data from the Longitudinal Experiences And Perceptions (LEAP) wave of TEDS data collection. A subset of 1,773 of these families were then invited to participate in the LEAP-2 follow-up study approximately 9 months later, which included measures of sensitivity and life events. Selection for LEAP-2 consisted of families in which one or both twins had scored highly in measures of psychotic experiences in the LEAP study (1420 pairs) and ‘control’ families in which neither twin had high scores in these measures (355 pairs). Of the families invited, 1,471 (83%) returned the LEAP-2 booklets. Families with LEAP-2 data were more ethnically diverse and included a higher proportion of female twins than those who were not invited to or did not complete the LEAP-2 study, but they did not differ on family socioeconomic characteristics or in the proportion of twins who reported a life event (Supplementary Table S1). Individuals who completed at least one of the measures of environmental sensitivity, anxiety sensitivity or reported life events in LEAP-2 formed the study sample (∼500 monozygotic twin pairs and ∼900 dizygotic twin pairs). In accordance with standard exclusion criteria for TEDS analyses, participants with severe medical disorders, who experienced severe perinatal complications, or with unknown demographic variables or zygosity were excluded (https://www.teds.ac.uk/datadictionary/exclusions.htm). This resulted in a final sample of 2,939 individuals (59% female) with an average age of 17.1 years (SD = 0.9).

### Ethical approval

Ethical approval for TEDS was provided by the King’s College London Ethics Committee (reference: PNM/09/10–104). Written informed consent was obtained prior to each wave of data collection from parents and from twins themselves from age 16 onwards.

### Measures

#### Environmental sensitivity

Environmental sensitivity was assessed with the 12-item Highly Sensitive Child (HSC) scale, developed to capture the typical behaviours and experiences of sensitive children and adolescents (Pluess et al., 2018).The HSC scale assesses three domains of environmental sensitivity. Ease of excitation relates to becoming mentally overwhelmed by contextual stimuli (e.g. “I find it unpleasant to have a lot going on at once”). Aesthetic sensitivity assesses awareness of details and aesthetic appreciation (e.g. “I notice when small things have changed in my environment”). Low sensory threshold is characterised by unpleasant reactivity to sensory stimuli (e.g. “Loud noises make me feel uncomfortable”). Participants are asked to rate the extent to which each statement describes them, on a Likert scale ranging from ‘Not at all’ (1) to ‘Extremely’ (7). The internal consistency of the scale in TEDS is α = 0.81 for the full scale, and α = 0.64-0.81 for the domain subscales (Assary et al., 2021), in line with previous studies (Booth et al., 2015; Smolewska et al., 2006). Responses were summed to give total environmental sensitivity scores, with higher scores representing higher levels of sensitivity. A copy of the full questionnaire is given in Supplementary Table S2. The full LEAP-2 study booklet is accessible through the TEDS data dictionary (http://www.teds.ac.uk/datadictionary; Rimfeld et al., 2019).

#### Anxiety sensitivity

Anxiety sensitivity was assessed using the Children’s Anxiety Sensitivity Index (CASI), which captures fear of anxiety sensations (Silverman et al., 1991). The CASI is an 18-item self-report questionnaire that asks participants whether statements such as ‘Unusual feelings in my body scare me’ are ‘Not true’ (0), ‘Quite true’ (1) or ‘Very true’ (2) over the last six months. The internal consistency of the CASI in TEDS is α = .93 (Eley et al., 2007). Responses were summed to give total anxiety sensitivity scores, with higher scores representing higher levels of sensitivity. A copy of the full questionnaire is given in Supplementary Table S3.

#### Reported life events

Reported life events were assessed using a reduced version of the Coddington Life Events Scale (Coddington, 1972), which comprised the 20 items most relevant to adolescents. Participants were asked to self-report whether they had experienced any of the events in the past six months and, if the event had occurred, whether they found the experience ‘Very unpleasant’, ‘Moderately unpleasant’, ‘Neither unpleasant nor pleasant’, ‘Moderately pleasant’ or ‘Very pleasant’. Of the 20 items, 8 are considered to be ‘family-wide’, for example ‘Marital separation of my parents’, as they relate to the general family environment. The remaining 12 items are considered to be ‘twin-specific’, for example ‘Failing an important exam’ (Supplementary Table S5). Both types of events were retained in this analysis, as family-wide experiences can be differentially perceived by individual siblings (Daniels et al., 1985) and the correlation between twins on the total number of reported family-wide events was only moderate (ρ = 0.43). A copy of the full questionnaire is given in Supplementary Table S4 and a breakdown of the proportion of responses for each event in Supplementary Table S5.

This measure was used to derive three variables (Supplementary Table S6). First, the total number of reported life events was established by converting each item to a binary response. To do this, all responses indicating the experience of an event (‘Yes’, ‘Very unpleasant’, ‘Moderately unpleasant’, ‘Neither unpleasant nor pleasant’, ‘Moderately pleasant’ and ‘Very pleasant’) were collapsed into one category representing ‘Present’ (1), while the response option ‘No’ represented ‘Absent’ (0). The number of present events were summed to calculate the total number of reported life events. Therefore, scores for the total number of life events ranged from 0 to 20. This variable was used in Models 1 and 3. For Model 2, variables representing valence ratings of life events reported as being positive and negative were created. Positive ratings of events were calculated by summing ‘Moderately pleasant’ (1) and ‘Very pleasant’ (2) responses. Negative ratings of events were calculated by summing ‘Moderately unpleasant’ (1) and ‘Very unpleasant’ (2) responses. The responses ‘No’ and ‘Neither unpleasant nor pleasant’ were coded as 0 for both variables. In this way, whether an event is positive or negative was determined individually for each twin, based upon their own ratings. As such, items contributing to positive ratings of events for one individual could contribute to negative ratings for another. Scores for positive and negative ratings of events ranged from 0 to 40.

### Analyses

#### Descriptive statistics

Descriptive statistics including means, standard deviation and skewness were assessed for all variables. Phenotypic correlations were estimated in the full sample and in males and females separately. Cross-twin correlations were estimated for each measure for monozygotic (MZ) and dizygotic (DZ) twins, to give an indication of genetic and environmental influences.

#### Genetic analyses

Using model fitting of twin data, the contribution of genetic and environmental influences to individual differences in a trait can be estimated (Knopik et al., 2017). MZ twin pairs share 100% of their genes, whilst DZ twins share on average 50%. If it is assumed that both types of twin pairs share their environments to similar extents, a greater degree of similarity in a trait between MZ twin pairs compared to DZ twin pairs reflects genetic influences (A). When correlations between DZ twin pairs are more than half of those between MZ twin pairs, similarity reflects shared environmental influences (C). Differences between MZ twin pairs are used to infer non-shared environmental influences (E) which also include any measurement error. In multivariate models, these principles can be applied to estimate the aetiology of the associations between multiple traits, using cross-twin cross-trait correlations. Higher cross-twin cross-trait correlations for MZ twins compared to DZ twins indicates that covariance between two traits can be attributed to genetic influences.

To prevent inflation of the correlation between twins, variables were adjusted for age and sex prior to model fitting, by regressing each variable on both covariates and using the residuals in subsequent analyses (McGue & Bouchard, 1984). For measures with skewness scores greater than 1, residuals were mapped onto a normal distribution using the rank-based van der Waerden’s transformation. Genetic model fitting was conducted within R (R Core Team, 2017) using the structural equation modeling package OpenMx (Neale et al., 2016). To account for variations in sample sizes across the three measures, all models were fitted to the raw data using Full Information Maximum Likelihood (FIML), which enables the estimation of variance components and confidence intervals in analyses with missing data, assuming that data is missing at random (Newman, 2014).

Univariate analyses were first conducted to assess the genetic, shared environmental and non-shared environmental influences on each variable. Multivariate genetic analyses were then conducted in three stages. To address our first aim, the Cholesky decomposition interpreted as a multivariate correlated factors solution was used to examine the shared genetic and environmental influences between environmental sensitivity, anxiety sensitivity and the total number of reported life events (Model 1).

For our second aim, this model was extended by separating reported life events into those rated by twins as having negative and positive valences respectively, to explore their differential associations with anxiety sensitivity and environmental sensitivity (Model 2).

Finally, a variation of the trivariate Cholesky model was used to investigate the proportion of genetic and environmental influences on reported life events shared with environmental sensitivity and anxiety sensitivity (Model 3). In this model, the associations between environmental sensitivity and anxiety sensitivity were represented using a correlated factors solution so no direction of effect between these measures was inferred. In contrast, the association between the sensitivity measures and life events was interpreted as a Cholesky decomposition path, allowing the genetic and environmental effects on sensitivity to be accounted for in the estimation of the effects on life events.

To facilitate multivariate genetic model fitting, means, variances and within-person correlations were constrained to be equal across zygosity and birth order, and cross-twin correlations were constrained to be symmetrical. To test equality of means, variances and correlations, constrained phenotypic models in which these constraints were specified were compared to corresponding saturated models in which these parameters were freely estimated. Variances and covariances were passed into A, C and E components (ACE models). For models which included small and non-significant estimates of C, we assessed the fit of the more parsimonious AE submodel. Model comparisons were based on likelihood ratio testing using χ^2^ values and degrees of freedom (Kline, 2015).

## Results

### Descriptive statistics

Descriptive statistics for all variables in the study sample are presented in Table 1. After adjusting for age and sex, skewness greater than 1 persisted for all variables except environmental sensitivity, hence these variables were transformed. Phenotypic correlations for the full sample are given in Table 2 and were similar for males and females (Supplementary Table S7). Within-pair correlations for the transformed variables were higher for MZ twins than DZ twins, indicating the influence of genetic factors. Of note, the DZ correlations were less than half of the MZ correlations for anxiety sensitivity, environmental sensitivity and negative ratings of life events, suggesting that estimates of A may include some non-additive genetic effects. We retained the ACE specification based on previous evidence that models specifying non-additive genetic effects (D) did not fit the data better than when C influences were specified for either anxiety sensitivity (Zavos, Gregory, et al., 2012) or environmental sensitivity (Assary et al., 2021).

**Table 1.** Descriptive statistics and intraclass twin correlations (n = 2939)

| Measure | Raw data |  | Variables regressed on age and sex |  | Cross-twin correlations for transformed variables |  |
| --- | --- | --- | --- | --- | --- | --- |
| | N | Mean (SD) | Skew before transformation | Skew after transformation | $r_{MZ}$ (95% CI) | $r_{DZ}$ (95% CI) |
| Anxiety sensitivity | 2862 | 8.3 (6.4) | 1.08 | 0.00 | .50 (.44, .56) | .15 (.09, .22) |
| Environmental sensitivity | 2799 | 35.8 (11.3) | 0.01 | - | .49 (.42, .55) | .22 (.15, .28) |
| Number of life events | 2584 | 1.7 (1.6) | 2.49 | 0.00 | .50 (.43, .56) | .28 (.21, .35) |
| Negative ratings of life events | 2471 | 1.2 (1.6) | 2.14 | 0.00 | .47 (.40, .54) | .21 (.14, .28) |
| Positive ratings of life events | 2471 | 1.2 (1.5) | 1.29 | 0.00 | .40 (.32, .48) | .27 (.20, .33) |

**Table 2.** Multivariate results: standardised variance components for each measure, cross-twin cross-trait correlations and phenotypic correlations between measures of sensitivity and life events with proportions of variance explained by A and E

| Measures | Standardised variance components (95% CI) | Associations with sensitivity measures (95% CI) |  |  |  |  |  |
| --- | --- | --- | --- | --- | --- | --- | --- |
|  |  | Anxiety sensitivity |  |  | Environmental sensitivity |  |  |
| | | Cross-twin cross-trait correlations | Phenotypic correlations | Proportion of $r_{ph}$ explained by A and E | Cross-twin cross-trait correlations | Phenotypic correlations | Proportion of $r_{ph}$ explained by A and E |
| Anxiety sensitivity | $h^2$: .47<br>(.40 - .52)<br>$e^2$: .53<br>(.48 - .60) | - | | - | - | - | |
| Environmental sensitivity | $h^2$: .48<br>(.42 - .54)<br>$e^2$: .52<br>(.46 - .58) | $r_{MZ}$: .37<br>(.32 - .42)<br>$r_{DZ}$: .14<br>(.08 - .19) | $r_{ph}$: .59<br>(.57 - .62) | A: 60%<br>(52 - 67%)<br>E: 40%<br>(33 - 48%) | - | - | |
| Number of life events | $h^2$: .51<br>(.45 - .57)<br>$e^2$: .49<br>(.43 - .55) | $r_{MZ}$: .16<br>(.10 - .21)<br>$r_{DZ}$: .04<br>(-.01 - .09) | $r_{ph}$: .21<br>(.17 - .25) | A: 70%<br>(49 - 89%)<br>E: 30%<br>(11 - 51%) | $r_{MZ}$: .12<br>(.07 - .17)<br>$r_{DZ}$: .02<br>(-.03 - .07) | $r_{ph}$: .15<br>(.11 - .19) | A: 69%<br>(41 - 96%)<br>E: 31%<br>(4 - 59%) |
| Negative ratings of life events | $h^2$: .46<br>(.39 - .52)<br>$e^2$: .54<br>(.48 - .61) | $r_{MZ}$: .18<br>(.12 - .23)<br>$r_{DZ}$: .03<br>(-.02 - .08) | $r_{ph}$: .20<br>(.16 - .24) | A: 75%<br>(53 - 97%)<br>E: 25%<br>(3 - 47%) | $r_{MZ}$: .12<br>(.06 - .18)<br>$r_{DZ}$: .01<br>(-.04 - .06) | $r_{ph}$: .14<br>(.10 - .18) | A: 72%<br>(38 - 104%)<br>E: 28%<br>(-4 - 62%) |
| Positive ratings of life events | $h^2$: .43<br>(.36 - .49)<br>$e^2$: .57<br>(.51 - .64) | $r_{MZ}$: .06<br>(.01 - .12)<br>$r_{DZ}$: .00<br>(-.05 - .05) | $r_{ph}$: .09<br>(.05 - .13) | A: 59%<br>(-1 - 111%)<br>E: 41%<br>(-11 - 101%) | $r_{MZ}$: .06<br>(.01 - .12)<br>$r_{DZ}$: .03<br>(-.02 - .08) | $r_{ph}$: .10<br>(.05 - .14) | A: 67%<br>(16 - 116%)<br>E: 33%<br>(-16 - 85%) |

### Univariate models

Univariate model fitting results are presented in Supplementary Table S8, and were consistent with previous estimations (Assary et al., 2021; Waszczuk et al., 2015; Wootton et al., 2017).

### Multivariate models

Comparison of saturated and constrained models indicated that the assumptions of equality of means and variances were met (*p* = .128-.249). Across ACE models, C estimates were generally small (<9%) and non-significant. For all models, dropping the C parameters did not result in significant worsening of fit (Models 1 and 3: χ^2^(6)=2.585057, *p*=.859, Model 2: χ^2^(10)=6.644232, *p*=.759). Hence, AE models are presented. The full ACE models are presented in Supplementary Figures S1 and S2. Model fit statistics and comparisons are given in Supplementary Tables S9 and S10.

The results of the multivariate models are presented below and summarised in Table 2. Standardised A and E influences are shown as h^2^ and e^2^ for each of the variables. To illustrate associations between measures of sensitivity and life events variables, cross-twin cross-trait correlations are given for MZ (r_MZ_) and DZ (r_DZ_) twin pairs, and within-person phenotypic correlations (r_ph_) are presented with the standardised proportions of the phenotypic associations accounted for by additive genetic (A) and unique environmental (E) influences.

#### Model 1

The Cholesky decomposition, represented as a multivariate correlated factors solution, was used to examine the genetic and environmental relationship between anxiety sensitivity, environmental sensitivity and the number of reported life events (Fig. 1).

**Figure 1.**
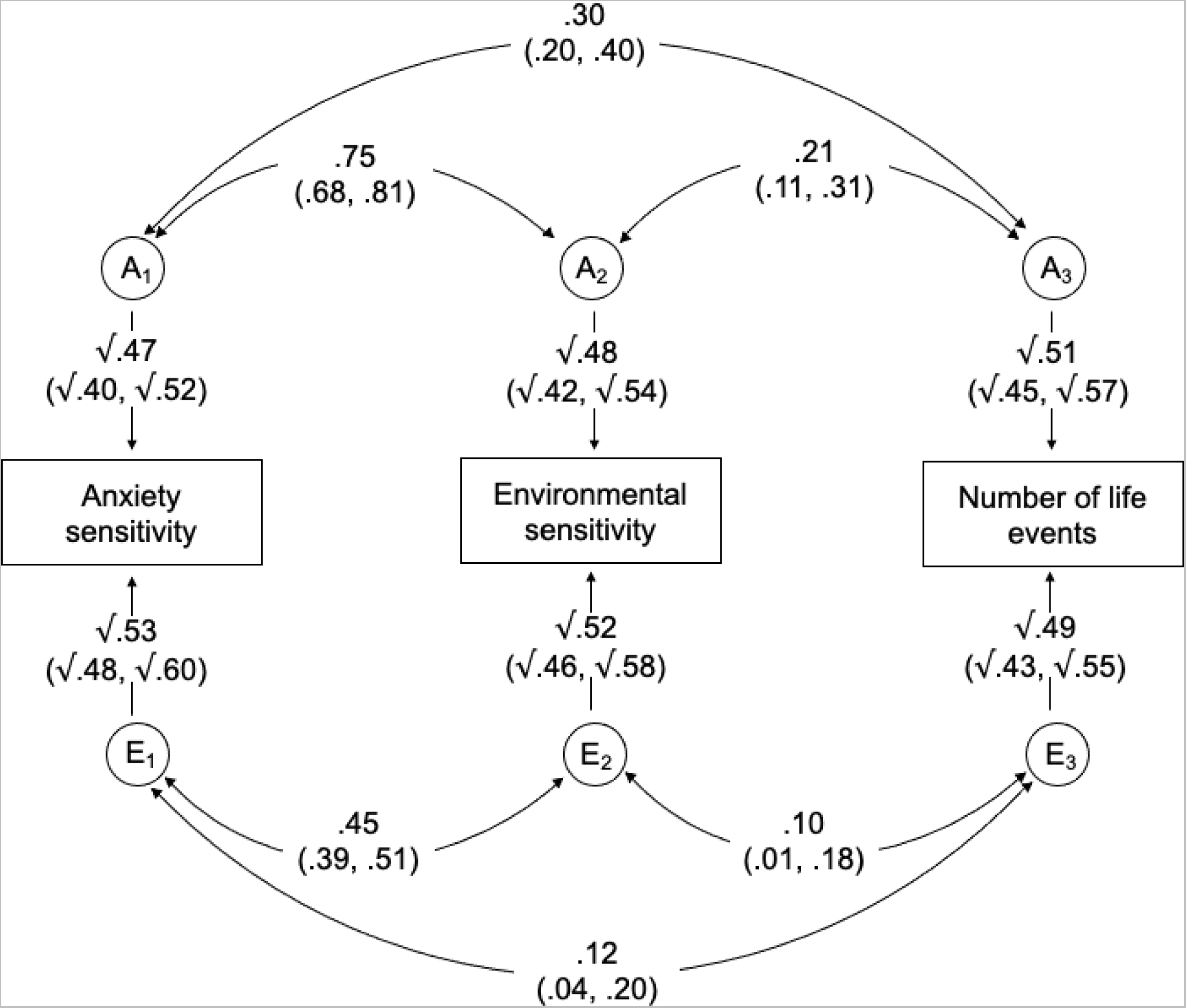
Correlated factors solution of the multivariate Cholesky decomposition for anxiety sensitivity, environmental sensitivity and number of life events. A_1-3_ and E_1-3_ represent the respective additive genetic and non-shared environmental influences (95% CIs). Curved paths show the correlations between the A and E factors for each measure (95% CIs).

Approximately half of the variance in each trait was explained by genetic influence (h^2^ = .47-.51), with the other half accounted for by non-shared environmental influences (e^2^ = .49-.53). Both anxiety sensitivity and environmental sensitivity showed moderate genetic and small non-shared environmental correlations with the number of reported life events (r_A_ = .30 and .21, r_E_ = .12 and .10, respectively; Fig. 1). Genetic influences accounted for 70% of the phenotypic correlation between anxiety sensitivity and life events, and 69% for environmental sensitivity and life events (Table 2).

#### Model 2

The multivariate correlated factors solution of Model 1 was extended to examine the shared genetic and environmental influences between the two measures of sensitivity and life events rated as having negative and positive valence (Fig. 2).

**Figure 2.**
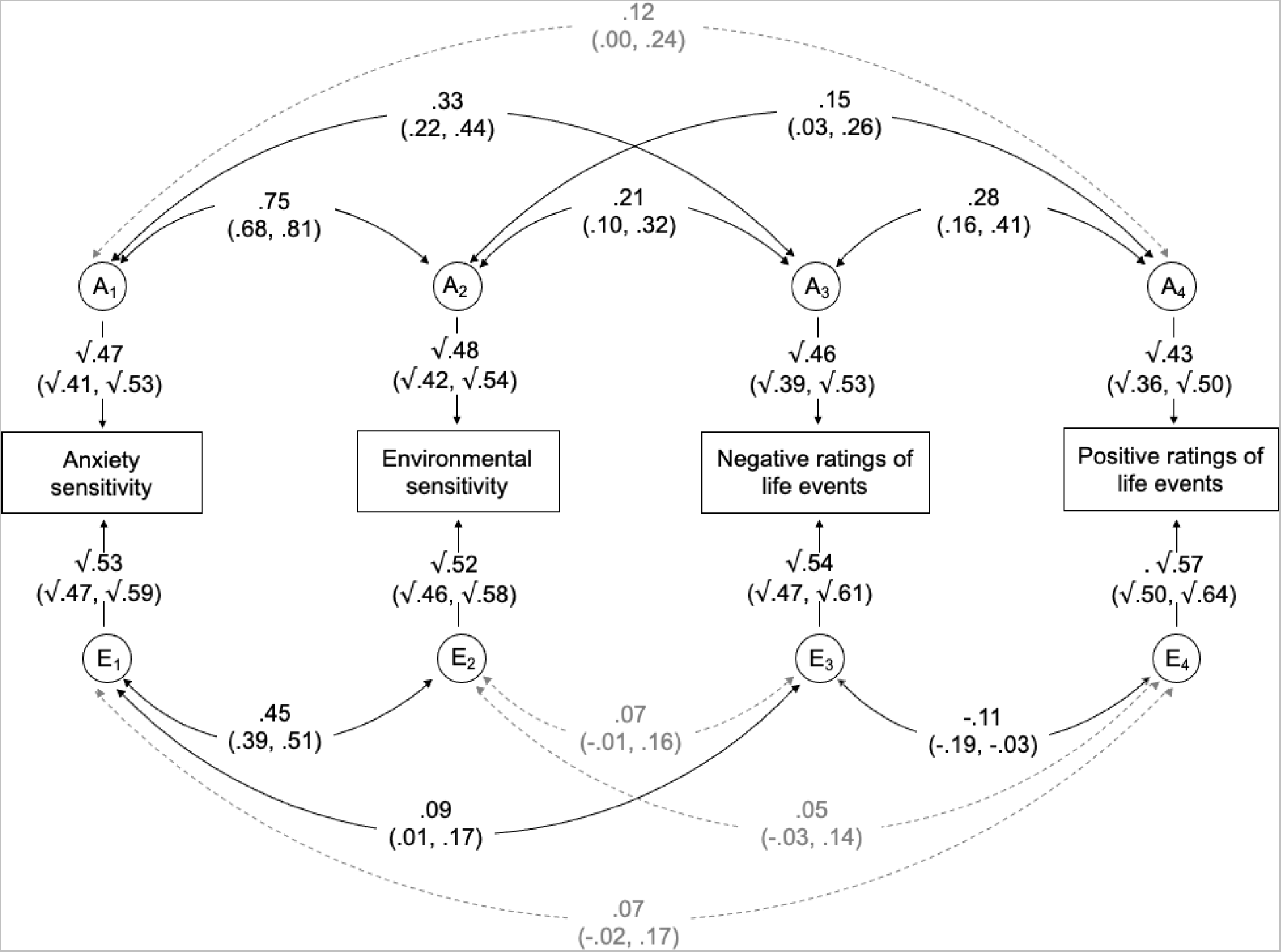
Correlated factors solution of the multivariate Cholesky decomposition for anxiety sensitivity, environmental sensitivity, negative ratings of life events and positive ratings of life events. A_1-4_ and E_1-4_ represent the respective additive genetic and non-shared environmental influences (95% CIs). Curved paths show the correlations between the A and E factors for each measure (95% CIs).

Negative ratings of life events showed moderate genetic correlations with both anxiety sensitivity and environmental sensitivity (rA = .33 and .21, respectively), and a small non-shared environmental correlation with anxiety sensitivity (r_c_ = .09). For positive ratings of life events, genetic and environmental correlations with both measures were lower and only the genetic correlation with environmental sensitivity was significant (r_A_ = .15).

#### Model 3

A variation of the trivariate Cholesky model of anxiety sensitivity, environmental sensitivity and number of reported life events was used to investigate the proportion of genetic and environmental influences on reported life events shared with anxiety sensitivity and environmental sensitivity (Fig. 3). In this model, the concurrent associations between anxiety sensitivity and environmental sensitivity are represented using a correlated factors solution and the associations between the sensitivity measures and life events are interpreted as Cholesky decomposition paths.

**Figure 3.**
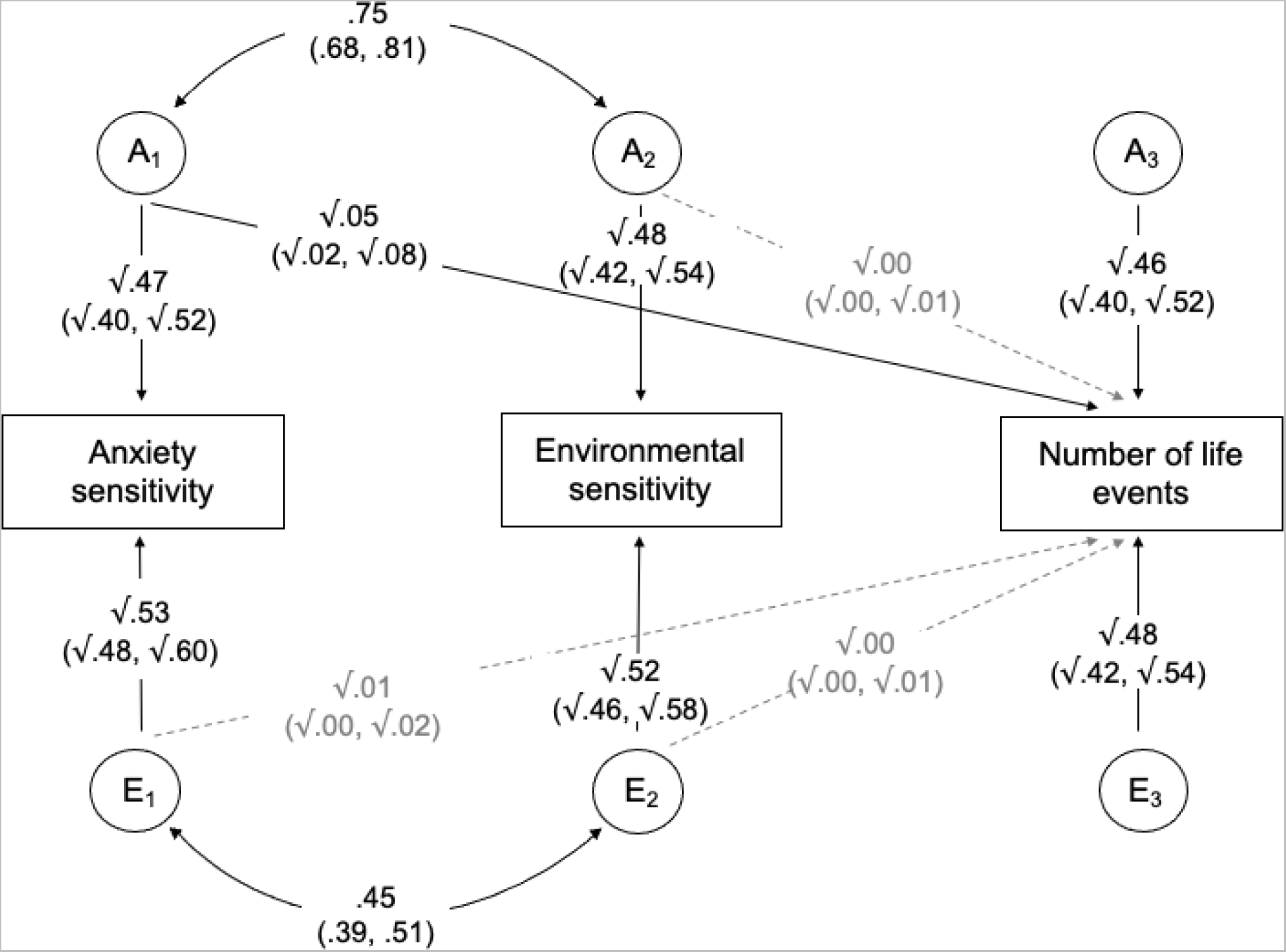
Unique genetic and environmental influences on number of life events, over and above those shared with anxiety sensitivity and environmental sensitivity. A_1-3_ and E_1-3_ represent the respective additive genetic and non-shared environmental influences (95% CIs). Curved paths show the correlation between the A and E factors for anxiety sensitivity and environmental sensitivity (95% CIs).

Genetic effects on anxiety sensitivity had a small but significant influence on reported life events (√.05 = .22). After controlling for the genetic influences shared with anxiety sensitivity, there were no additional unique genetic influences on reported life events from environmental sensitivity. After accounting for the genetic effects on both sensitivity measures, unique genetic influences on the number of reported life events accounted for 46% of the variance, compared to 51% when anxiety sensitivity and environmental sensitivity were not adjusted for (Model 1).

Hence, 90% (95% CIs 84-96%) of the genetic influences on reported life events were independent of the influences on anxiety sensitivity and environmental sensitivity (46%/51% × 100). Therefore, 10% (95% CIs 4-16%) of the genetic influences on reported life events were accounted for by the genetic influences on these measures. Almost all, 98% (95% CIs 96-100%), of the non-shared environmental influences on reported life events were independent of the non-shared environmental influences on the sensitivity measures (48%/49% × 100).

## Discussion

The aim of this study was to explore the shared genetic and environmental influences on sensitivity and reported life events in adolescence. We investigated sensitivity biases related to how individuals process the contextual aspects of their environment (environmental sensitivity) and how they interpret their own physical and emotional responses (anxiety sensitivity). The majority of the associations between all measures were explained by shared genetic influences (59%-75%), with the remainder explained by non-shared environmental influences (25%-41%). Environmental sensitivity showed comparable genetic correlations with both negative and positive ratings of life events (r_A_ = .21 and .15), whereas anxiety sensitivity only showed a significant genetic correlation with negative ratings of life events (r_A_ = .33). Approximately 10% of the genetic influences on reported life events were accounted for by the genetic influences on anxiety sensitivity and environmental sensitivity.

### Genetic influences on environmental and anxiety sensitivity

In line with previous research, anxiety sensitivity and environmental sensitivity showed moderate heritability, with the remaining variance explained by non-shared environmental influences (Assary et al., 2021; Zavos et al., 2010). The high genetic contribution to the covariance between these measures suggests that the phenotypic association between these two different aspects of sensitivity is driven by shared genetic influences. These findings are suggestive of pleiotropy, whereby the same genes influence both traits. For environmental sensitivity, previous studies have found evidence for a multi-dimensional genetic model, with genetic influences consisting of those related to general sensitivity, heightened response to negative stimuli, and sensitivity to more positive aspects of the environment (Assary et al., 2021). As anxiety sensitivity also captures negative interpretation of physical and emotional responses, it is logical that a large proportion of its genetic influences are shared with the genetic influences on overall environmental sensitivity. Therefore, the present findings suggest that many of the same genetic factors that influence overall response to external stimuli also influence the interpretation of these physical and emotional responses as harmful.

### Genetic influences on life events

As expected, life events also displayed moderate heritability (Kendler & Baker, 2007). Covariance between life events and the sensitivity traits was predominantly genetically driven, with genetic effects accounting for 59-75% of the associations between life events and anxiety sensitivity, and 67-72% of the association between life events and environmental sensitivity. Genetic influences on environmental measures such as life events can be indicative of gene– environment correlation, whereby an individual’s genetic factors influence the environments that they are exposed to, either passively through parents, or actively or evocatively through genetically-influenced behaviours (Jaffee & Price, 2007). In this context, gene-environment correlation could occur through genetic influences on sensitivity creating a tendency for individuals to seek out or elicit certain environments that increase the likelihood of experiencing major life events (Zavos, Wong, et al., 2012). If this mechanism were at play, we would expect that individuals with greater genetic loading for sensitivity would also report greater exposure to life events.

An alternative plausible explanation for genetic overlap is that the genetic factors that influence sensitivity are not correlated with *exposure* to life events, but rather they influence an individual’s subjective experience. As life events were assessed using self-report, this measure is also likely to capture individual differences in the interpretation or impact of events, whether and how they are recalled, and willingness to disclose personal experiences. These aspects of self-reporting life events may share a genetic basis with sensitivity. This explanation is consistent with the relatively low phenotypic correlations between sensitivity and life events (r_ph_=.09-.21), which indicates that expression of these measures share limited overall variance. This may imply that while sensitivity contributes to the subjective experience of an event, as captured by self-reports, it cannot fully account for differences in exposure to environmental risk. However, it should be noted that the small phenotypic correlations between sensitivity and life events result in large confidence intervals around the proportions of covariance attributable to genetic factors, indicating that these estimates should be interpreted with caution.

### Differential associations with negative and positive appraisals of life events

The differential pattern of associations between the two types of sensitivity with negative and positive ratings of life events is consistent with the theoretical basis of these measures. The finding that environmental sensitivity was significantly genetically correlated with both positive and negative appraisals may suggest that higher genetic loading for this form of sensitivity is associated with greater appraisal of both adverse events as negative, and pleasant events as positive, which may attenuate the effects of such events on outcomes for these individuals. This explanation is in line with previous research that found more sensitive individuals are affected more negatively by adverse contexts but also more positively in response to positive exposures, compared to those who are generally less sensitive to both (Pluess et al., 2020). On the other hand, the underlying genetic liability of anxiety sensitivity is more relevant to the appraisal of events as negative and the tendency to perceive anxiety responses to these events as being harmful. This corresponds to previous findings that anxiety sensitivity moderates the association between stressful life events and later internalising symptoms (McLaughlin & Hatzenbuehler, 2009). This suggests that while sensitivity to contextual aspects of the environment may be important to the experience of both negative and positive events, sensitivity to one’s own anxiety responses may play a greater role in the perception of an environment as adverse.

### Sensitivity accounts for a proportion of the heritability of life events

Lastly, our findings suggest that a proportion of the heritable component of life events is captured by genetic influences on sensitivity. In accordance with the high genetic correlation between the sensitivity measures, genetic overlap with reported life events was driven by the shared genetic influences on sensitivity, with no independent contribution of environmental sensitivity once influences shared with anxiety sensitivity were accounted for. This indicates that differences in how individuals interpret the contextual aspects of or responses to their environments may be one mechanism through which genetic variation influences the experience of life events. Nonetheless, a substantial proportion of the genetic influences on life events were independent of these sensitivity measures. Other efforts to elucidate the heritable component of environmental experiences indicate that some of these remaining influences are likely to be accounted for by genetic influences on psychopathology and other related constructs, such as personality factors and cognitive biases, which may influence either exposure to or the subjective experience of environmental events (ter Kuile et al., 2022; Peel et al., 2022). Together, this knowledge contributes to understanding of what is captured by the heritable component of measures of environmental experiences, and the potential ways in which these experiences may be associated with risk for poor outcomes.

### Strengths and limitations

There are a number of strengths in this study. First, it demonstrates the utility of the twin design in exploring genetic overlap for complex traits. There is considerable variation in the conceptualisation and assessment of life events across different research studies. This variation means it is difficult to obtain the large, homogeneous samples required for molecular genetic analyses. Where sufficient data are available there are limitations surrounding sample ascertainment. For example, reports of stressful or traumatic life events are often collected in large-scale mental health studies, enabling investigation into the genetic variants associated with adversity in the context of disorder (Clarke et al., 2019; Power et al., 2013). However, studies focussed on mental health are typically enriched for affected individuals, therefore, results are unlikely to reflect the general population (Power et al., 2013). Furthermore, there is a scarcity of measures capturing the experience of positive events, despite these also being relevant to mental health and wellbeing (Luhmann et al., 2012). Although the specific genetic variants underlying sensitivity cannot be detected through the twin design, this analysis provides initial evidence that a proportion of the genetic underpinnings of self-reported life events are shared with sensitivity in a sample of adolescents that is representative of the UK population (Haworth et al., 2013). Additionally, to our knowledge, this is the first investigation of differential patterns of genetic and environmental influences between sensitivity and negative and positive appraisals of life events. This enabled the investigation of how the theoretical assumptions of environmental and anxiety sensitivity translate at the genetic level.

However, a number of limitations should also be considered. First, dependent and independent life events were grouped together, although there is some evidence indicating that the former display greater genetic influences (Kendler & Baker, 2007). This decision was made due to the difficulty in categorising events based on the assumed influence of an individual’s behaviour, which is likely to vary widely throughout the sample. For example, ‘Being hospitalized for illness or injury’ could be either a dependent or independent event. Secondly, the classification of negative and positive events was determined individually based on each twin’s appraisals. This was decided with the aim of capturing individual perception rather than assessing exposure to predetermined negative/positive events, and because appraisals for each event were distributed across the full scale of responses (Supplementary Table S5). However, this approach does limit the conclusions that can be drawn about influences on the reporting of specific types of events. Additionally, all measures were collected at the same time point, hence, data are cross-sectional and causality should not be assumed. Although interpretations are primarily given in the direction of sensitivity influencing life events, it is also possible that the experience of life events influences the development of sensitivity (Zavos, Wong, et al., 2012). As data on sensitivity and positive life events were only collected in the LEAP-2 assessment, we were not able to utilise longitudinal data to assess the direction of association in these analyses. Additionally, it was not possible to assess whether the characteristics of this subsample, primarily the overrepresentation of individuals who scored highly on measures of psychotic experiences, might have impacted on self-reporting of these measures. However, heritability estimates for anxiety sensitivity in this study were comparable to estimates in adolescent twins from an alternative large UK twin sample (Zavos, Gregory, et al., 2012). Finally, the twin design has some inherent limitations, including the assumption that MZ and DZ twin pairs share their environments to the same degree (Rijsdijk & Sham, 2002)

### Implications

This study contributes to knowledge of what is captured by the heritable component of self-reported life events. Due to their ease of application and low cost, self-reported measures are becoming an increasingly popular means of assessing environmental experiences in large research studies. As such, it is important to investigate the traits that influence their reporting, in order to increase understanding of the aspects of experience that are captured by these measures. Our findings indicate that sensitivity biases are among these relevant traits, displaying shared genetic propensity with reporting of life events, and appraisal of events as negative or positive. This work reinforces the importance of using genetically-sensitive designs when investigating life events as an environmental risk factor. This may be especially important to consider when examining associations between reported life events and outcomes related to sensitivity in adolescence, including depression (Waszczuk et al., 2015), anxiety (Zavos, Rijsdijk, et al., 2012) and personality traits (Assary et al., 2021), as genetic influences are common across these phenotypes. Furthermore, it demonstrates the need for nuanced interpretation of self-reported life events as a measure that captures elements of both exposure and subjective experience.

The finding that sensitivity biases are among the heritable factors that comprise the genetic component of life events is consistent with growing evidence that the subjective perception of the environment plays an important role in its impact on the individual (Danese & Widom, 2020). Further knowledge of how sensitivity relates to the experience and outcomes of life events could provide novel avenues for mental health interventions in those with high genetic propensity for sensitivity traits (Assary, Krebs & Eley, 2022). Currently, the majority of research investigating the heritable basis of sensitivity has focussed on the expression of these traits in childhood and adolescence. As the genetic influences on many related constructs, including life events (Johnson et al., 2013), are found to increase throughout development, a key avenue for future research is the investigation of these relationships at later stages of development.

## Data Availability

The data that support the findings of this study came from the Twins Early Development Study (TEDS). Eligible researchers can apply for access to the TEDS data at https://www.teds.ac.uk/researchers/teds-data-access-policy.

https://osf.io/haud4/

## Acknowledgements

The authors gratefully acknowledge the on-going contribution of the participants in the TEDS and their families.

