## Supplementary for "A multivariate genetic analysis of environmental sensitivity, anxiety sensitivity and reported life events in adolescents"

**Contents**

**Tables** 3

Table S1. Comparison of Twins Early Development Study (TEDS) families at age 16 with and without data from the LEAP-2 booklet (n = 9917) 3

Table S2. Highly Sensitive Child Questionnaire (Pluess et al., 2018) 4

Table S3. Children's Anxiety Sensitivity Index (Silverman et al., 2013) 5

Table S4. Coddington Life Events Scale (Coddington, 1972) 6

Table S5. Classification of twin-specific and family-wide events from the Coddington Life Events Scale (Coddington, 1972) and proportion of responses for each event (n = 2939) 8

Table S6. Response coding for three variables created from the Coddington Life Events Scale (Coddington, 1972) 10

Table S7. Phenotypic Pearson’s correlations between all variables for the full sample and split by sex 11

Table S8. Univariate model fitting results 12

Table S9. Multivariate model fitting results with 95% confidence intervals 12

Cross-twin cross-trait correlations (below diagonal), phenotypic correlations with proportion of variance explained by A and E (above diagonal) and standardised variance components (on diagonal). Brackets indicate 95% CIs. 12

Table S10. Model fit statistics for Models 1 and 3 (including environmental sensitivity, anxiety sensitivity and number of reported life events) 13

Table S11. Model fit statistics for Model 2 (including environmental sensitivity, anxiety sensitivity, negative ratings of life events and positive ratings of life events) 14

**Figures** 15

Figure S1. Full ACE model of environmental sensitivity, anxiety sensitivity, number of life events 15

Figure S2. Full ACE model of environmental sensitivity, anxiety sensitivity, negative ratings of life events and positive ratings of life events 16

### Tables

**Table S1.** Comparison of Twins Early Development Study (TEDS) families at age 16 with and without data from the LEAP-2 booklet (n = 9917)

| | Families with LEAP-2 data | Families without LEAP-2 data | Comparison of families with and without LEAP-2 data |
|---|---|---|---|
| Number of individuals | 2943 | 6974 | 9917 |
| White ethnicity | 92.5% | 93.9% | $\chi^2(1) = 5.2404$, $p = .022$, $V = .02$* |
| Mothers with A-levels or higher | 24.0% | 22.7% | $\chi^2(1) = 0.0001$, $p = .992$, $V = .00$ |
| Fathers with A-levels or higher | 19.8% | 20.1% | $\chi^2(1) = 0.9345$, $p = .334$, $V = .02$ |
| Female | 57.8% | 54.6% | $\chi^2(1) = 8.1743$, $p = .004$, $V = .03$* |
| MZ | 35.2% | 36.6% | $\chi^2(1) = 1.7094$, $p = .191$, $V = .01$ |
| Reported at least one life event in LEAP Study | 75.0% | 76.1% | $\chi^2(1) = 2.1104$, $p = .146$, $V = .02$ |

*Notes:* * = difference significant at $p < 0.05$

**Table S2.** Highly Sensitive Child Questionnaire (Pluess et al., 2018)

**For each of the following statements, please select the point on the scale that you feel is most appropriate in describing you.**

| | Not at all | | | Moderately | | | Extremely |
|---|---|---|---|---|---|---|---|
| | 1 | 2 | 3 | 4 | 5 | 6 | 7 |
| 1. I notice when small things have changed in my environment | | | | | | | |
| 2. Loud noises make me feel uncomfortable | | | | | | | |
| 3. I love nice smells | | | | | | | |
| 4. I get nervous when I have to do a lot in little time | | | | | | | |
| 5. Some music can make me really happy | | | | | | | |
| 6. I am annoyed when people try to get me to do too many things at once | | | | | | | |
| 7. I don't like watching TV programmes that have a lot of violence in them | | | | | | | |
| 8. I find it unpleasant to have a lot going on at once | | | | | | | |
| 9. I don't like it when things change in my life | | | | | | | |
| 10. I love nice tastes | | | | | | | |
| 11. I don't like loud noises | | | | | | | |
| 12. When someone observes me, I get nervous. This makes me perform worse than normal | | | | | | | |

**Table S3.** Children's Anxiety Sensitivity Index (Silverman et al., 2013)

**How true are the following statements when you think about your feelings over the last six months?**

| | Not true | Quite true | Very true |
|---|---|---|---|
| | 0 | 1 | 2 |
| 1. I don't want other people to know when I feel afraid | | | |
| 2. When I cannot keep my mind on my schoolwork, I worry that I might be going crazy | | | |
| 3. It scares me when I feel "shaky" | | | |
| 4. It scares me when I feel like I am going to faint | | | |
| 5. It is important for me to stay in control of my feelings | | | |
| 6. It scares me when my heart beats fast | | | |
| 7. I feel embarrassed when my stomach rumbles or makes noise | | | |
| 8. It scares me when I feel like I am going to throw up | | | |
| 9. When I notice that my heart is beating fast, I worry that there might be something wrong with me | | | |
| 10. It scares me when I have trouble getting my breath | | | |
| 11. When my stomach hurts, I worry that I might be really ill | | | |
| 12. It scares me when I cannot concentrate on my schoolwork | | | |
| 13. Others my age can tell when I feel shaky | | | |
| 14. Unusual feelings in my body scare me | | | |
| 15. When I am afraid, I worry that I might be crazy | | | |
| 16. I get scared when I feel nervous | | | |
| 17. I don't like to let my feelings show | | | |
| 18. Funny feelings in my body scare me | | | |

**Table S4.** Coddington Life Events Scale (Coddington, 1972)

**Here is a list of events that might have happened to you recently. Please put a tick in either the 'No' or 'Yes' box to say whether the event has happened in the past six months. If you answer 'yes' then please indicate what it was like, choosing one of the options given, ranging from 'very unpleasant' to 'very pleasant'.**

In the past six months, I have experienced...

| | Yes | No | Very unpleasant | Moderately unpleasant | Neither unpleasant nor pleasant | Moderately pleasant | Very pleasant |
|---|---|---|---|---|---|---|---|
| 1. The loss of a job by my father or mother | | | | | | | |
| 2. Marital separation of my parents | | | | | | | |
| 3. Becoming involved with drugs | | | | | | | |
| 4. The death of a close friend or relative | | | | | | | |
| 5. Being hospitalized for illness or injury | | | | | | | |
| 6. Being sent away from home | | | | | | | |
| 7. Breaking up with a boyfriend/girlfriend | | | | | | | |
| 8. The hospitalization of my brother or sister | | | | | | | |
| 9. Suspension from school/college | | | | | | | |
| 10. Failing an important exam | | | | | | | |
| 11. Remarriage of a parent to a stepparent | | | | | | | |
| 12. Hospitalization of a parent | | | | | | | |

| |
|---|
| 13. Being responsible for a road accident |
| 14. A major decrease in parental income |
| 15. Getting pregnant or fathering a pregnancy |
| 16. Outstanding personal achievement |
| 17. Decrease in number of arguments between parents |
| 18. Becoming a member of a church |
| 19. Beginning to date |
| 20. Moving to a new school or college |

**Table S5.** Classification of twin-specific and family-wide events from the Coddington Life Events Scale (Coddington, 1972) and proportion of responses for each event (n = 2939)

| | Number of responses (%) | | | | | | |
|---|---|---|---|---|---|---|---|
| | No | Very unpleasant | Moderately unpleasant | Neither unpleasant nor pleasant | Moderately pleasant | Very pleasant | NA |
| **Family-wide events** | | | | | | | |
| The loss of a job by my father or mother | 2704 (92.0%) | 34 (1.2%) | 76 (2.6%) | 52 (1.8%) | 3 (0.1%) | 2 (0.1%) | 68 (2.3%) |
| Marital separation of my parents | 2820 (96.0%) | 28 (1.0%) | 20 (0.7%) | 13 (0.4%) | 0 | 1 (0.0%) | 57 (1.9%) |
| The death of a close friend or relative | 2479 (84.4%) | 270 (9.2%) | 111 (3.8%) | 19 (0.7%) | 5 (0.2%) | 3 (0.1%) | 52 (1.8%) |
| The hospitalization of my brother or sister | 2705 (92.0%) | 37 (1.3%) | 53 (1.8%) | 20 (0.7%) | 1 (0.0%) | 0 | 123 (4.2%) |
| Remarriage of a parent to a stepparent | 2847 (96.9%) | 8 (0.3%) | 8 (0.3%) | 4 (0.1%) | 11 (0.4%) | 6 (0.2%) | 55 (1.9%) |
| Hospitalization of a parent | 2721 (92.6%) | 85 (2.9%) | 65 (2.2%) | 19 (0.6%) | 1 (0.0%) | 2 (0.1%) | 46 (1.6%) |
| A major decrease in parental income | 2666 (90.7%) | 79 (2.7%) | 102 (3.5%) | 38 (1.3%) | 1 (0.0%) | 0 | 53 (1.8%) |
| Decrease in number of arguments between parents | 2277 (77.5%) | 25 (0.9%) | 21 (0.7%) | 40 (1.4%) | 221 (7.5%) | 246 (8.4%) | 109 (3.7%) |
| **Twin-specific events** | | | | | | | |
| Becoming involved with drugs | 2751 (93.6%) | 6 (0.2%) | 9 (0.3%) | 42 (1.4%) | 51 (1.7%) | 12 (0.4%) | 68 (2.3%) |
| Being hospitalized for illness or injury | 2736 (93.1%) | 46 (1.6%) | 62 (2.1%) | 33 (1.1%) | 5 (0.2%) | 3 (0.1%) | 54 (1.8%) |
| Being sent away from home | 2800 (95.3%) | 6 (0.2%) | 20 (0.7%) | 14 (0.5%) | 23 (0.8%) | 17 (0.6%) | 59 (2.0%) |

| | | | | | | | |
|---|---|---|---|---|---|---|---|
| Breaking up with a boyfriend/girlfriend | 2463 (83.8%) | 191 (6.5%) | 145 (4.9%) | 49 (1.7%) | 16 (0.5%) | 11 (0.4%) | 64 (2.2%) |
| Suspension from school/college | 2850 (97.0%) | 9 (0.3%) | 15 (0.5%) | 12 (0.4%) | 3 (0.1%) | 6 (0.2%) | 44 (1.5%) |
| Failing an important exam | 2297 (78.2%) | 344 (11.7%) | 184 (6.3%) | 46 (1.6%) | 2 (0.1%) | 4 (0.1%) | 62 (2.1%) |
| Being responsible for a road accident | 2878 (97.9%) | 4 (0.1%) | 6 (0.2%) | 3 (0.1%) | 0 | 2 (0.1%) | 46 (1.6%) |
| Getting pregnant or fathering a pregnancy | 2863 (97.4%) | 10 (0.4%) | 4 (0.1%) | 7 (0.2%) | 3 (0.1%) | 3 (0.1%) | 49 (1.7%) |
| Outstanding personal achievement | 1864 (63.4%) | 44 (1.5%) | 15 (0.5%) | 29 (1.0%) | 252 (8.6%) | 662 (22.5%) | 73 (2.5%) |
| Becoming a member of a church | 2839 (96.6%) | 6 (0.2%) | 2 (0.1%) | 6 (0.2%) | 14 (0.5%) | 14 (0.5%) | 58 (2.0%) |
| Beginning to date | 2449 (83.3%) | 14 (0.5%) | 18 (0.6%) | 33 (1.1%) | 130 (4.4%) | 228 (7.8%) | 67 (2.3%) |
| Moving to a new school or college | 2548 (86.7%) | 18 (0.6%) | 50 (1.7%) | 83 (2.8%) | 109 (3.7%) | 78 (2.7%) | 53 (1.8%) |

**Table S6.** Response coding for three variables created from the Coddington Life Events Scale (Coddington, 1972)

| Variable | Response coding | | | | | | |
|---|---|---|---|---|---|---|---|
| | No | Yes | Very unpleasant | Moderately unpleasant | Neither unpleasant nor pleasant | Moderately pleasant | Very pleasant |
| **Total number of life events** | 0 | 1 | 1 | 1 | 1 | 1 | 1 |
| **Positive ratings of events** | 0 | 0 | 0 | 0 | 0 | 1 | 2 |
| **Negative ratings of events** | 0 | 0 | 2 | 1 | 0 | 0 | 0 |

**Table S7.** Phenotypic Pearson's correlations between all variables for the full sample and split by sex

| | **Full sample (n = 2939)** | **Males (n = 1239)** | **Females (n = 1700)** |
|---|---|---|---|
| Anxiety sensitivity - Environmental sensitivity | .59 (.57-.62) | .60 (.56-.63) | .60 (.57-.63) |
| Anxiety sensitivity - Number of life events | .20 (.16-.24) | .23 (.17-.28) | .20 (.15-.25) |
| Anxiety sensitivity - Negative ratings of life events | .19 (.15-.23) | .24 (.18-.29) | .16 (.11-.21) |
| Anxiety sensitivity - Positive ratings of life events | .09 (.05-.13) | .12 (.06-.18) | .09 (.04-.15) |
| Environmental sensitivity - Number of life events | .15 (.11-.19) | .13 (.07-.19) | .16 (.11-.21) |
| Environmental sensitivity - Negative ratings of life events | .13 (.09-.17) | .15 (.08-.21) | .14 (.08-.19) |
| Environmental sensitivity - Positive ratings of life events | .10 (.06-.14) | .09 (.03-.15) | .10 (.05-.16) |
| Negative ratings of life event - Positive ratings of life events | .07 (.03-.11) | .10 (.04-.16) | .12 (.07-.17) |

**Table S8.** Univariate model fitting results

| Measure | ACE variance components (95% CI) | | |
|---|---|---|---|
| | **A** | **C** | **E** |
| Anxiety sensitivity | .46 (.39 - .52) | .00 (.00 - .04) | .54 (.48 - .60) |
| Environmental sensitivity | .48 (.35 - .54) | .00 (.00 - .09) | .52 (.46 - .58) |
| Number of life events | .45 (.26 - .56) | .05 (.00 - .19) | .50 (.44 - .57) |
| Negative ratings of life events | .46 (.31 - .52) | .00 (.00 - .10) | .54 (.48 - .61) |
| Positive ratings of life events | .29 (.09 - .49) | .12 (.00 - .27) | .69 (.52 - .67) |

**Table S9.** Model fit statistics for Models 1 and 3 (including environmental sensitivity, anxiety sensitivity and number of reported life events)

| **Base model** | **Comparison** | **−2LL** | **df** | **AIC** | **Δ-2LL** | **Δdf** | **ΔAIC** | **p** |
|---|---|---|---|---|---|---|---|---|
| Saturated | | 34973.03 | 8161 | 35081.03 | | | | |
| Saturated | Constrained | 35011.12 | 8194 | 35053.12 | 38.09378 | 33 | -27.91 | .249 |
| Saturated | ACE | 35019.47 | 8194 | 35061.47 | 46.4404 | 33 | -19.56 | .060 |
| Constrained | ACE | 35019.47 | 8194 | 35061.47 | 8.346612 | 0 | 8.35 | - |
| Saturated | **AE** | **35022.05** | **8200** | **35052.05** | 49.02545 | 39 | -28.98 | .130 |
| Constrained | **AE** | **35022.05** | **8200** | **35052.05** | 10.93167 | 6 | -1.07 | .091 |
| ACE | **AE** | **35022.05** | **8200** | **35052.05** | 2.585057 | 6 | -9.42 | .859 |

*Notes*: Saturated = model with the maximum number of parameters to get a baseline index of fit; Constrained = submodel of Saturated model with means and variances constrained to be equal across twin order and zygosity, to test the assumptions of the twin design; ACE = model with A, C and E parameters representing genetic, shared environment and non-shared environment influences; AE = submodel where C parameters are fixed to 0; -2LL = minus twice the log likelihood; df = degrees of freedom; AIC Akaike's information criterion; Δ-2LL = difference in -2LL value; Δ df= difference in degrees of freedom; ΔAIC= difference in AIC value; p = p-value of Δ-2LL. The ACE model and was a good fit to the data as compared to the saturated model ($p$ = .060). Bold typeface indicates final model selected.

**Table S10.** Model fit statistics for Model 2 (including environmental sensitivity, anxiety sensitivity, negative ratings of life events and positive ratings of life events)

| Base model | Comparison | −2LL | df | AIC | Δ-2LL | Δdf | ΔAIC | p |
|---|---|---|---|---|---|---|---|---|
| Saturated | | 41508.10 | 10476 | 41684.10 | | | | |
| Saturated | Constrained | 41574.04 | 10530 | 41642.04 | 65.94461 | 54 | -42.06 | .128 |
| Saturated | ACE | 41586.45 | 10530 | 41654.45 | 78.35471 | 54 | -29.65 | .017 |
| Constrained | ACE | 41586.45 | 10530 | 41654.45 | 12.41011 | 0 | 12.41 | - |
| Saturated | **AE** | **41593.10** | **10540** | **41641.10** | 84.99895 | 64 | -43.00 | .041 |
| Constrained | **AE** | **41593.10** | **10540** | **41641.10** | 19.05434 | 10 | -0.94 | .040 |
| ACE | **AE** | **41593.10** | **10540** | **41641.10** | 6.644232 | 10 | -13.35 | .759 |

*Notes*: Saturated = model with the maximum number of parameters to get a baseline index of fit; Constrained = submodel of Saturated model with means and variances constrained to be equal across twin order and zygosity, to test the assumptions of the twin design; ACE = model with A, C and E parameters representing genetic, shared environment and non-shared environment influences; AE = submodel where C parameters are fixed to 0; -2LL = minus twice the log likelihood; df = degrees of freedom; AIC Akaike's information criterion; Δ-2LL = difference in -2LL value; Δ df= difference in degrees of freedom; ΔAIC= difference in AIC value; p = p-value of Δ-2LL. The ACE model did not fit the data as well as the saturated model (*p* = .017). This can occur in studies with large sample sizes as minimal variance differences between groups can be highly statistically significant. The significant difference between the ACE and saturated model may reflect differences in the variance component influences indicated by the data and as specified in the genetic model. Bold typeface indicates final model selected.

**Figures**

**Figure S1.** Full ACE model of environmental sensitivity, anxiety sensitivity, number of life events

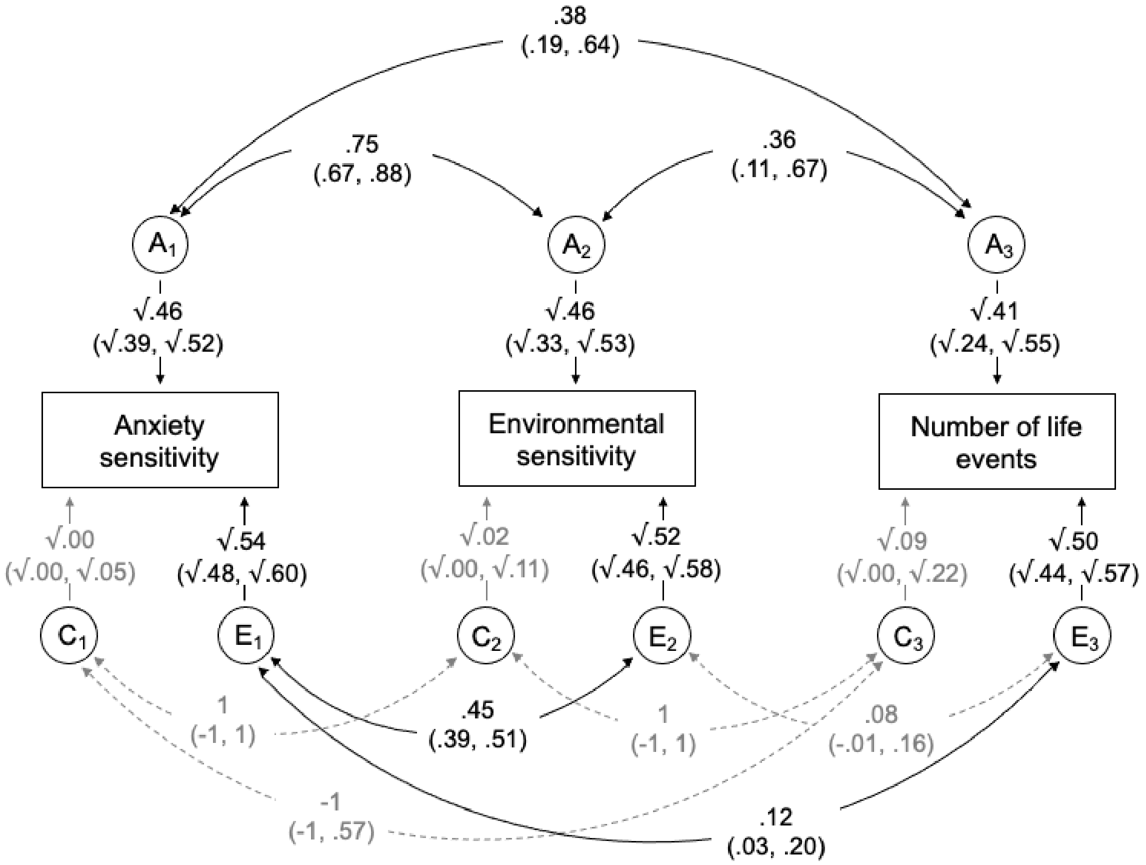

*Figure S1.* Correlated factors solution of the multivariate Cholesky decomposition for anxiety sensitivity, environmental sensitivity and number of life events. $A_{1\text{-}3}$, $C_{1\text{-}3}$ and $E_{1\text{-}3}$ represent the respective additive genetic, shared environmental, and non-shared environmental influences (95% CIs). Curved paths show the correlations between the A, C and E factors for each measure (95% CIs).

**Figure S2.** Full ACE model of environmental sensitivity, anxiety sensitivity, negative ratings of life events and positive ratings of life events

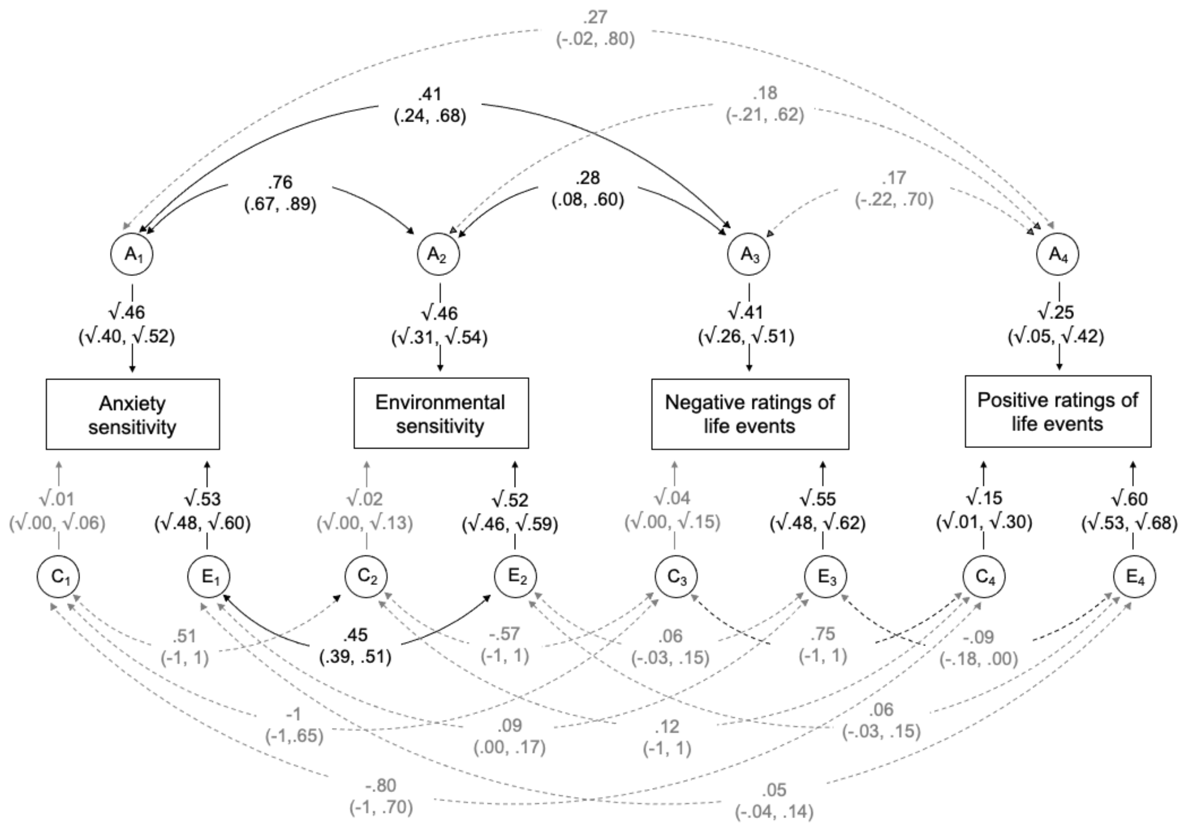

*Figure S2.* Correlated factors solution of the multivariate Cholesky decomposition for anxiety sensitivity, environmental sensitivity, negative ratings of life events and positive ratings of life events. $A_{1\text{-}4}$, $C_{1\text{-}4}$ and $E_{1\text{-}4}$ represent the respective additive genetic, shared environmental and non-shared environmental influences (95% CIs). Curved paths show the correlations between the A, C and E factors for each measure (95% CIs).
